# Rational Design and Effectiveness Analysis of Pediatric Medication Pathways under DRG/DIP Payment Models

**DOI:** 10.64898/2026.01.10.26343843

**Authors:** Yi Pengfei, Zhu Chunhui, Wan Muyuan, Wang Bin, Peng Dan, Tang Xiaolin

## Abstract

**Objective:** To evaluate the impact of implementing pediatric medication pathways on operational management under the DRG/DIP payment model in a specialized children’s hospital, using DRG as a case study.

**Methods:** All medical records with DRG codes DT13 and GW15 from 2023 (pre-implementation control group) and 2024 (post-implementation observation group) were included. A systematic comparative analysis of clinical data was performed to assess the effects on hospital operational metrics.

**Results:** The observation group exhibited a significantly lower average cost per case (DT13: 17.82%; GW15: 26.05%) and a higher medical insurance payment margin (DT13: ¥186,500; GW15: ¥89,000) compared to the control group. The examination cost proportion and total hospitalization expenses decreased significantly (P < 0.05), whereas the drug cost proportion showed a non-significant decreasing trend (P > 0.05). Regarding efficiency, the average length of stay and number of drug varieties were significantly reduced (P < 0.05). Quality indicators, including antibiotic usage rate, antibiotic use intensity, and adverse drug reaction incidence, were significantly improved (P < 0.05), alongside an increased rational prescription review rate. Furthermore, the number of high-cost cases decreased while low-cost cases increased for both disease groups post-implementation, with all case distributions remaining within clinically acceptable bounds.

**Conclusion:** Pediatric medication pathways represent an effective strategy for balancing cost-containment with the unique clinical demands of pediatric care within the DRG/DIP framework. They provide a practical reference for precision management in pediatric hospitals and empirical evidence to inform pediatric-sensitive medical insurance payment policies.

## 1. Introduction

The Diagnosis-Related Groups (DRG) system is a patient classification framework that categorizes patients based on disease severity, treatment complexity, and resource consumption. Each group is assigned a fixed payment rate, and hospitals are reimbursed by medical insurance based on this predetermined standard corresponding to the patient’s DRG category ^[1]^. The Diagnosis-Intervention Packet (DIP) model employs big data analytics to define payment units grounded in the combination of “diagnosis + treatment modality” ^[2]^. In contrast to the traditional fee-for-service model, both DRG and DIP are designed to incentivize medical institutions to optimize clinical processes, control costs, and enhance the efficiency of healthcare resource utilization ^[1, 2]^.

Among medical specialties, pediatrics presents distinct challenges attributable to the unique characteristics of its patient population. Children are not merely small adults; their disease spectrum, pathophysiological features, pharmacokinetics, and pharmacodynamics differ substantially from those of adults ^[3]^. These physiological and developmental distinctions necessitate more tailored and individualized design of pediatric clinical pathways and medication treatment plans ^[3]^. However, prevailing DRG/DIP grouping systems are primarily derived from adult disease profiles, often lacking sufficient incorporation of pediatric specificities, which can lead to adaptation challenges during implementation ^[4]^.

Evidence suggests that drug costs represent a higher proportion of total treatment expenditures in pediatrics compared to adult medicine ^[4]^. This financial pressure is compounded by the pronounced shortage of child-specific medication formulations, further exacerbating the operational strain on pediatric departments under DRG/DIP ^[4]^.

Clinical medication pathways constitute a critical tool for bridging healthcare quality and cost-control objectives, holding particular relevance within the DRG/DIP context ^[5]^. By standardizing diagnostic and therapeutic processes and optimizing medication strategies, these pathways can facilitate cost-control objectives without compromising healthcare quality ^[5]^. In pediatrics, a scientifically rational pathway design is imperative, as it impacts not only institutional operational efficiency but also directly influences treatment safety and long-term outcomes for pediatric patients. However, systematic research focused on the design of pediatric medication pathways within the DRG/DIP framework remains limited, particularly empirical evidence elucidating how to balance cost-control with pediatric-specific clinical needs ^[6]^.

This study aims to perform an in-depth analysis of the design principles and implementation strategies for pediatric clinical medication pathways under the DRG/DIP payment model. Employing a combination of DRG theoretical frameworks and empirical analysis, this study seeks to identify optimized pathways suitable for the pediatric healthcare context in China ^[7]^. The findings are intended to provide a reference for children’s hospitals to enhance pharmaceutical care under healthcare payment reform and to offer evidence for policymakers to develop more pediatric-adaptive payment policies. Ultimately, this research strives to contribute to a multi-faceted benefit: safeguarding children’s health rights, ensuring the sustainable development of children’s hospitals, and promoting the efficient utilization of medical insurance funds ^[8]^.

## 2. Materials and Methods

### 2.1 Study Population and Design

A retrospective cohort study was conducted.Medical records from patients reimbursed under the DRG/DIP payment model in our hospital in 2023, prior to the establishment of pediatric medication pathways, were designated as the control group. Records from 2024, following pathway implementation, were assigned to the observation group. Cases with DRG codes DT13 (Otitis Media and Upper Respiratory Infection, with Complications or Comorbidities) and GW15 (Esophagitis, Gastroenteritis, without Complications or Comorbidities) were selected for analysis.

### 2.2. Inclusion and exclusion criteriaControl

Control Group 1 (DT13): 449 cases (273 males, 176 females); age range: 1 month 3 days to 15 years; mean age: 6.0 ± 3.2 years.

- Observation Group 1 (DT13): 523 cases (316 males, 207 females); age range: 1 month 14 days to 13 years 9 months; mean age: 5.6 ± 3.2 years.
- Control Group 2 (GW15): 345 cases (213 males, 132 females); age range: 1 month 14 days to 15 years; mean age: 6.3 ± 4.1 years.
- Observation Group 2 (GW15): 368 cases (224 males, 144 females); age range: 1 month 9 days to 16 years 4 months; mean age: 8.4 ± 5.1 years.

No statistically significant differences were observed in baseline characteristics between the control and observation groups for either disease group(P > 0.05), confirming comparability. Inclusion criteria were:(1) Age ≤ 18 years; (2) Primary diagnosis of either DT13 or GW15; (3) Complete medical record data.

Exclusion criteria were:(1) Incomplete medical record data; (2) Length of stay > 30 days; (3) Hospitalization cost < ¥10.

### 2.3 Method: Pediatric Medication Pathway Design

The design of the pediatric medication pathways was guided by a child-centered philosophy.This is critical as children, unlike adults, are in a rapid stage of growth and development. Consequently, treatment plans must consider not only the management of the acute condition but also potential impacts on long-term development. Therefore, pathway design transcended a purely disease-focused perspective, adopting a holistic, whole-process health management model [9]. Within the DRG/DIP framework, this child-centered philosophy was operationalized by extending pathway coverage to include post-discharge medication guidance, thereby aiming to prevent treatment interruptions or reduced efficacy associated with shortened hospital stays ^[10]^.

Both the control and observation groups were managed under the hospital’s standard DRG/DIP payment model.The observation group additionally received care guided by the newly designed pediatric clinical medication pathways for DT13 and GW15, which standardized medication selection and utilization for these conditions. Specific procedures included:

(1) Data Collection:Data including average cost per case, drug cost proportion, examination cost proportion, and length of stay were extracted from the hospital’s DRG system for both groups.
(2) DRG/DIP Grouping:Grouping was based on the National Healthcare Security Administration’s CHS-DRG grouping scheme and the local DIP catalog. DRG codes were uniformly calibrated by trained medical coders.
(3) Pediatric Medication Pathway Design:The pathways were developed based on current clinical guidelines. Key principles included: prioritizing medications listed in the medical insurance catalog; reducing the use of adjunctive therapies and inappropriate antibiotic combinations; and standardizing treatment regimens for the target diseases. These pathways can provide decision support prompts to physicians.

### 2.4. Statistical analysis

Statistical analyses were performed using SPSS(version 26.0). Continuous data are presented as mean ± standard deviation and were compared between groups using the independent samples t-test if normally distributed; otherwise, the Mann-Whitney U test was employed. Categorical data are presented as numbers (percentages) and were compared using the Chi-squared (χ²) test. A two-tailed P-value < 0.05 was defined as statistically significant.

### 2.5. Ethical statement

This study is a retrospective study, adhering to the principles of the Declaration of Helsinki and approved by the Ethics Committee of Jiangxi Provincial Children’s Hospital (JXEY-BG06A(1)). This study was also conducted in accordance with the approved clinical trial protocol (Registration Number: JXSETYYYXKY--20240130, First Registration Date: May 30, 2024, and exemption from informed consent was granted.

### 2.6 Outcome Measures

Outcome measures were categorized as follows:

- Economic: Average cost per case, drug cost proportion, examination cost proportion, consumables cost proportion, profit/loss amount, total hospitalization cost.
- Efficiency: Average length of stay, number of drug varieties per case.
- Quality: Antibiotic usage rate, antibiotic use intensity, rational rate of prescription reviews, incidence of adverse drug reactions.

### 2.7 The Importance of Pediatric Medication Pathways in Medical Services

The ongoing reform of the DRG/DIP payment method has demonstrated efficacy in curbing the irrational growth of medical expenditures. A comparative analysis of national data from 2018 (pre-reform) and 2022 (the inaugural year of the “Three-Year Action Plan”) revealed a notable decline in drug costs among 28 major disease categories tracked in the Health Statistics Yearbook [9, 10]. Specifically, 26 categories exhibited a reduced drug cost proportion, while 25 showed a decrease in the combined proportion of drugs and medical consumables. The most substantial reductions were observed in several oncology-related diseases. For instance, malignant neoplasms of the lung, stomach, and bladder witnessed decreases exceeding 10, 8, and 7 percentage points, respectively. This trend suggests that the DRG/DIP payment model has effectively contained the excessive growth of drug costs, even amidst the rapid launch and inclusion of numerous innovative anti-tumor agents in the medical insurance catalog ^[11, 12]^.

Despite these successes, the reform presents several challenges. From a cost perspective, a paradoxical trend emerged: although the drug cost proportion generally decreased, the per capita medical costs increased in 19 of the 28 disease categories, with an average annual growth rate of 2.4% ^[11, 12]^. This indicates that solely controlling the drug cost proportion does not automatically reduce total medical expenses. Institutions may adapt by increasing the volume and cost of examinations, treatments, and other services to compensate for lost drug revenue, a practice sometimes referred to as “volume expansion” to offset revenue loss.

Regarding clinical behavior, some hospitals, in an effort to minimize financial losses under the new payment scheme, have directly linked these losses to departmental and physician performance evaluations. This has led to the unintended consequence of incentivizing “accounting before doctoring,” where clinical decisions—including patient admission and treatment plans—are influenced by preemptive cost-benefit calculations. In more severe cases, cost control is circumvented through practices such as “upcoding” or “split hospitalizations.” Such practices can potentially compromise medical quality and detrimentally affect the patient experience.

It is noteworthy that, counter to the general trend, the drug cost proportion for certain conditions, such as acute myocardial infarction and aplastic anemia, increased by 1-3 percentage points. This phenomenon suggests that the cost-control leverage of DRG/DIP may be constrained for diseases that are highly pharmacotherapy-dependent and have limited alternative treatment options. This observation provides a crucial insight for the design of pediatric medication pathways: the unique clinical and pharmacological characteristics of different pediatric diseases must be thoroughly considered to avoid a simplistic, “one-size-fits-all” approach to cost control. In our pediatric specialty hospital setting, prior to pathway implementation, the management of DRG codes DT13 and GW15 was characterized by relatively high rates of antibiotic utilization and excessive use of non-core adjunctive therapies. The subsequent optimization of medication regimens through the establishment of structured pediatric medication pathways is detailed below (Table 1).

**Table 1.**
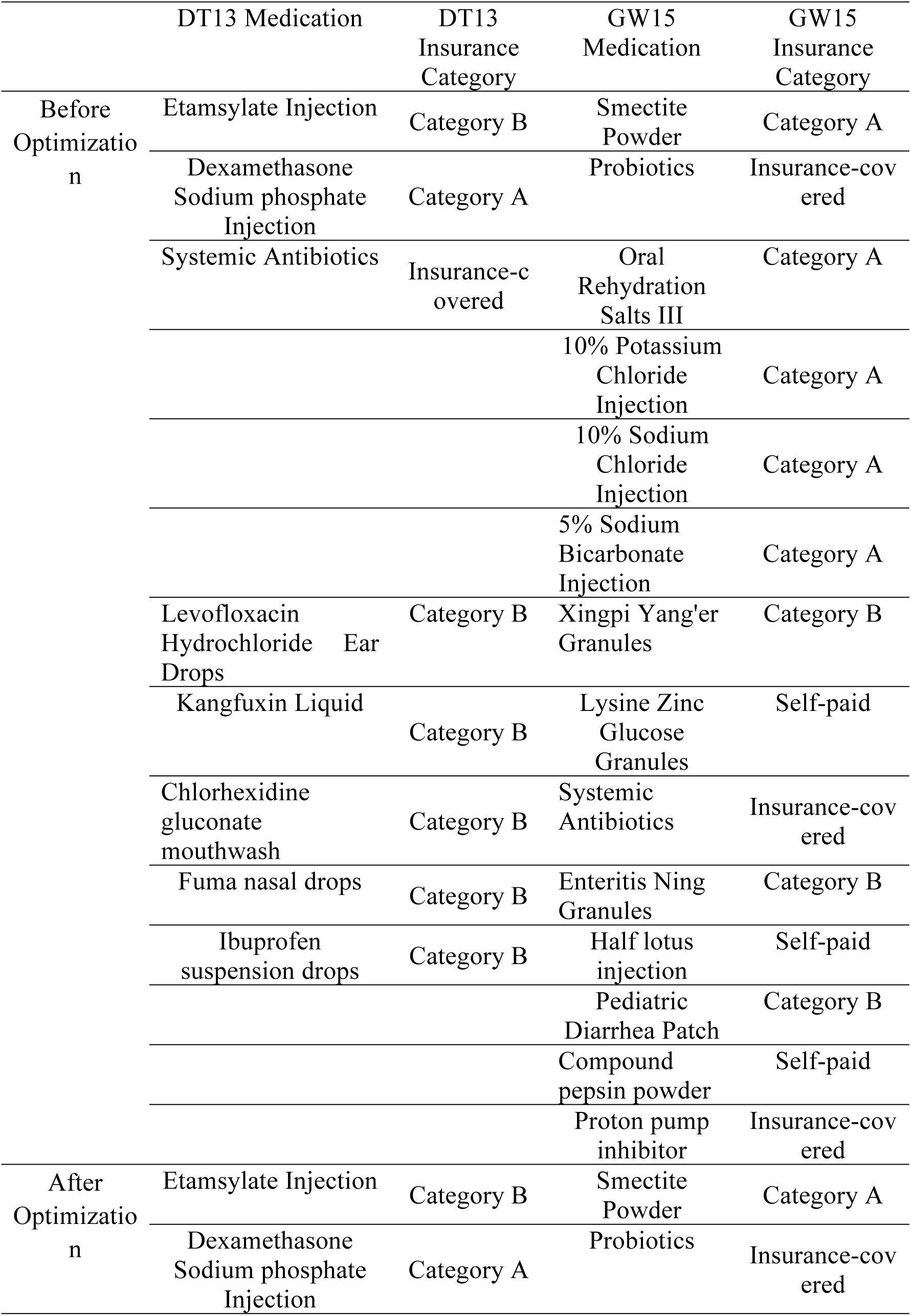
Optimization of medication regimens for DT13 and GW15 before and after pathway implementation.

## 3. Results

### 3.1 Comparison of Economic Indicators Post-Implementation (Details in Table 2-3)

The average cost of both observation groups was lower than that of the control group, and the difference was statistically significant (P<0.05); The profit and loss amounts of both observation groups were higher than those of the control group, and the difference was statistically significant (P<0.05); The above two sets of data indicate that the application of pediatric clinical drug routes has significant cost advantages for diseases.

**Table 2.** Comparison of Safety and Cost Indicators between Two Groups of Medical Services[n(%)]

|  |  |  |  |
| --- | --- | --- | --- |
| Budesonide Suspension for Inhalation | Category B | Oral Rehydration Salts III | Category A |
| Compound Ipratropium Bromide Solution for Inhalation | Category B | 10% Potassium Chloride Injection | Category A |
|  |  | 10% Sodium Chloride Injection | Category A |
|  |  | 5% Sodium Bicarbonate Injection | Category A |

**Table 3.** Comparison of Safety and Cost Indicators between Two Groups of Medical Services[n(%)]

| Group |  | Drug Cost Proportion | Examination Cost Proportion | Consumables Cost Proportion | Total Hospitalization Cost (¥000) |
| --- | --- | --- | --- | --- | --- |
| Control (DT13) | 1 | 19.18% | 55.92% | 1.86% | 1935.9 |
| Observation (DT13) | 1 | 16.49% | 43.86% | 4.06% | 1853.2 |
| Change 1 |  | -2.69% | -12.06% | 2.2% | -8.27 |
| $\chi^2/t$ | | 0.58 | 9.63 | 4.98 | 74.2 |
| P |  | >0.05 | <0.05 | <0.05 | <0.05 |
| Control (GW15) | 2 | 16.37% | 54.39% | 1.74% | 1853.5 |
| Observation (GW15) | 2 | 14.65% | 45.59% | 3.30% | 1462.1 |
| Change 2 |  | -1.72% | -8.80% | 1.56% | -39.14 |
| $\chi^2/t$ | | 0.37 | 4.86 | 2.16 | 209.48 |
| P |  | >0.05 | <0.05 | >0.05 | <0.05 |

There was a decrease in the proportion of medication between the two groups in 2024 and 2023, but the difference was not statistically significant (P>0.05). There was a statistically significant difference in the proportion of examination and total hospitalization expenses (P<0.05); There was a statistically significant difference in the proportion of consumption between group 1 (P<0.05), while there was no statistically significant difference between group 2 (P>0.05).

### 3.2 Comparison of Efficiency Indicators Post-Implementation (Details in Table 4)

After applying pediatric clinical drug routes, there was a statistically significant difference (P<0.05) in the average length of hospital stay and the selection of drug varieties between the two observation groups, both showing a certain degree of decrease. Among them, the length of hospital stay in group 1 was shortened by about 0.17 days, and group 2 was shortened by about 0.45 days.

**Table 4.** Comparison of two sets of efficiency indicators(x±s)

| Group | Average length of hospital stay (days) | Drug variety (species) |
| --- | --- | --- |
| Control 1 (DT13) | 3.86±0.57 | 10.25±0.82 |
| Observation 1 (DT13) | 3.69±0.39 | 6.71±0.75 |
| Reduction 1 | -4.40% | -34.54% |
| t | 5.48 | 70.38 |
| P | <0.05 | <0.05 |
| Control 2 (GW15) | 4.47±0.12 | 11.38±0.64 |
| Observation 2 (GW15) | 4.02±0.23 | 5.89±0.42 |
| Reduction2 | -10.07% | -48.24% |
| t | 32.37 | 135.9 |
| P | <0.05 | <0.05 |

### 3.3 Comparison of Quality Indicators Post-Implementation (Details in Table 5)

After applying pediatric clinical drug routes, there were significant statistical differences (P<0.05) in the usage rate, intensity of antibacterial drug use, reasonable rate of medical advice comments, and incidence of adverse drug reactions between the observation group and the control group, indicating a more rational use of antibacterial drugs

**Table 5.**
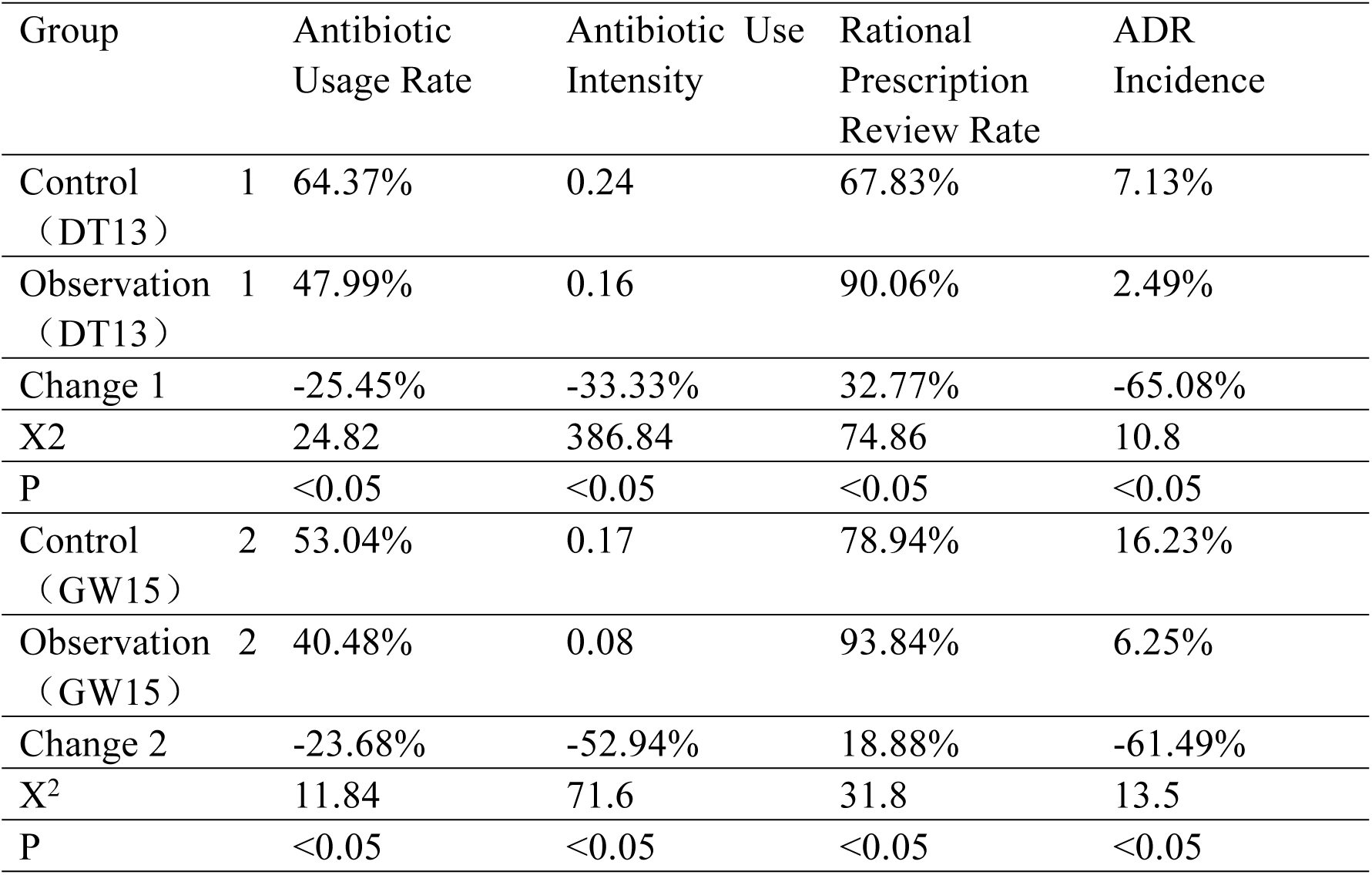
Comparison of two sets of quality indicators.

### 3.4 Comparison of Case Type Distribution Post-Implementation (Details in Table 6)

After adopting pediatric drug routes, the number of high magnification cases of DT13 and GW15 diseases decreased in 2024 compared to 2023, while the number of low magnification cases increased, but both remained within a reasonable range.

**Table 6.** Distribution of case types for two diseases.

| Disease | Year | Normal Cases | Low-Cost Cases | High-Cost Cases |
| --- | --- | --- | --- | --- |
| DT13 | 2023 | 308 (68.60%) | 58 (12.92%) | 83 (18.49%) |
|  | 2024 | 362 (69.22%) | 138 (26.39%) | 23 (4.40%) |
| GW15 | 2023 | 184 (53.33%) | 60 (17.39%) | 101 (29.28%) |
|  | 2024 | 206 (55.98%) | 111 (30.16%) | 51 (13.86%) |

## 4. Discussion

Under the DRG/DIP payment model, children’s specialized hospitals have achieved profits in some diseases by developing pediatric clinical drug routes. However, there are still many problems in the implementation process, and the inadequate adaptability of the grouping system is currently the main challenge faced by pediatrics ^[13]^. The existing DRG/DIP grouping is mainly designed based on the characteristics of adult diseases and treatment modes, with insufficient consideration for the specificity of pediatrics. Taking neonatal diseases as an example, low birth weight infants, premature infants, etc. need to be comprehensively evaluated for the severity of the condition and treatment intensity based on various factors such as gestational age, birth weight, and complications. However, the existing grouping system often cannot accurately reflect these complex factors. This has led to many pediatric cases being classified as “miscellaneous” or “other” groups, and the grouping weights or scores cannot truly reflect resource consumption, causing economic pressure on medical institutions when admitting critically ill children.

This study was conducted by pharmacists to optimize the clinical drug routes for different diseases. By comparing the clinical data of DRG codes DT13 (otitis media and upper respiratory tract infections, with complications or comorbidities) and GW15 (esophagitis and gastroenteritis, without complications or comorbidities) from a children’s specialized hospital in 2023 (without implementing pediatric drug routes) and 2024 (with implementing pediatric drug routes), the actual impact of pediatric drug routes on medical management under the DRG/DIP payment model was revealed. The two diseases mentioned in this study generally have a treatment cycle of 2 weeks, which may lead to no suitable grouping or grouping deviation due to the above reasons. Therefore, this study excluded two extreme phenomena (hospitalization time exceeding 30 days; hospitalization cost less than 10 yuan) to ensure their inclusion. The accuracy and rationality of the group.

### 4.1 Optimization of Medical Economic Outcomes

The implementation of pediatric medication pathways was associated with a significant reduction in the average cost per case(DT13: 17.82%; GW15: 26.05%) and a marked improvement in the medical insurance payment margin (DT13: +¥186,500; GW15: +¥89,000). These improvements can be largely attributed to the core pathway design principles, which emphasized the selection of medications within the medical insurance catalog and the reduction of non-essential adjunctive therapies. Although the reduction in drug cost proportion did not reach statistical significance (P > 0.05), a concurrent decrease was observed in the examination cost proportion. This broader reduction in costs may also have been influenced by external factors, such as policies promoting provincial mutual recognition of laboratory results and price reductions for specific diagnostic tests. The modest increase in consumables cost proportion had a negligible impact on the net reduction in total hospitalization costs. This pattern suggests that the pathways primarily achieved savings through the precise reduction of redundant drug expenditures, rather than through indiscriminate cost compression. Consequently, this approach did not trigger compensatory increases in other cost components, such as examinations or consumables, a phenomenon sometimes observed in response to single-minded cost-control measures.

### 4.2 Enhancement of Efficiency and Quality Metrics

The active involvement of pharmacists throughout the DRG/DIP disease management process—spanning pre-pathway design,intra-pathway monitoring, and post-pathway prescription review—was instrumental in achieving these results ^[10]^. Efficiency metrics demonstrated a shortened average length of stay (DT13: 0.17 days; GW15: 0.45 days) and a substantial reduction in the number of drug varieties per case (DT13: 34.54%; GW15: 48.24%). These improvements likely stem from the standardization of treatment processes facilitated by the pathways. The integration of pre-prescription review systems helped regulate drug selection, thereby minimizing unnecessary medication adjustments and potentially reducing prolonged hospitalization. Regarding quality indicators, significant reductions were observed in the antibiotic usage rate (DT13: 25.45%; GW15: 23.68%), antibiotic use intensity, and ADR incidence. Concurrently, the rational prescription review rate increased substantially (DT13: 32.77%; GW15: 18.88%). These findings indicate that the pathways were effective in enhancing medication safety for the pediatric population. The unique pharmacokinetic and pharmacodynamic profiles of pediatric patients necessitate heightened precision in drug therapy [3]. The pathways contributed to risk mitigation by restricting inappropriate antibiotic combinations and promoting the use of age-appropriate drug formulations ^[17]^.

### 4.3 Enhancing Adaptability to DRG/DIP Systems

Current DRG/DIP grouping systems,which are primarily derived from adult disease models, inadequately account for pediatric specificities. In this study, pathway implementation was followed by a decrease in high-cost cases and an increase in low-cost cases for both DT13 and GW15, with all case type distributions remaining within clinically acceptable ranges. This suggests that clinical pathways can improve the alignment of pediatric care with the DRG/DIP payment models by optimizing resource consumption patterns ^[4]^. For instance, for DT13, pathway intervention curbed the overuse of antibiotics and certain adjunctive therapies, thereby lowering resource consumption per case and reducing the incidence of high-cost cases (i.e., those exceeding payment standards). Simultaneously, by standardizing core treatments, the pathways helped maintain the quality of care for low-cost cases (i.e., those below payment standards), thus achieving a balance between cost control and quality assurance.

### 4.4 Limitations and Future Directions

The lack of statistical significance for certain economic indicators,such as the reduction in drug cost proportion, may be attributed to the sample size and the specific characteristics of the diseases studied. DT13 and GW15 are common conditions where baseline drug cost proportions may have already been relatively optimized, leaving limited room for drastic reduction. Consequently, the pathway optimization primarily involved “eliminating redundancies” rather than implementing “sharp cuts” ^[16, 17]^. Furthermore, the long-term effects of the pediatric pathways warrant further investigation. Key areas for future research include the impact of post-discharge follow-up on treatment continuity and the evaluation of long-term outcomes related to child growth and development. These aspects were beyond the scope of the present study.

## 5. Conclusion

In conclusion, the design and implementation of pediatric clinical medication pathways under the DRG/DIP payment model significantly improved the operational management of a specialized children’s hospital. The key findings are as follows:

- First, economic outcomes were optimized, evidenced by a significant reduction in the average cost per case and an increased medical insurance payment margin. This was achieved through the precise control of redundant drug expenditures, without triggering compensatory increases in costs for examinations or consumables.
- Second, medical efficiency was improved, as demonstrated by a shortened average length of stay and a reduced number of drug varieties used. These findings indicate enhanced resource utilization efficiency resulting from the standardization of treatment processes.
- Third, medical quality was assured. We observed reductions in irrational antibiotic use and the incidence of adverse drug reactions, alongside an improved rational prescription review rate. These improvements directly address the physiological particularities and medication safety requirements unique to the pediatric population.
- Finally, the pathways enhanced the adaptability of pediatric care to the DRG/DIP system. By adjusting the case cost structure—specifically, by reducing high-cost cases and stabilizing low-cost cases—the pathways facilitated a better fit for pediatric conditions within a payment system originally designed using adult data.

In summary, pediatric medication pathways represent an effective and practical tool for reconciling the dual objectives of cost-containment and addressing pediatric medical specificity within the DRG/DIP framework. They provide an actionable framework for the precision management of children’s hospitals and generate empirical evidence to inform the development of more pediatric-sensitive medical insurance payment policies. Future research should aim to expand the range of diseases covered by such pathways, extend the follow-up period to evaluate long-term sustainability, and refine the assessment of their impact on children’s long-term health outcomes ^[6, 7]^.

## Data Availability

All relevant data are within the manuscript and its Supporting Information files.

